# Mapping T cell activation and differentiation at single cell resolution in naive hosts infected with *Plasmodium vivax*

**DOI:** 10.1101/2021.03.22.21252810

**Authors:** Florian A. Bach, Diana Muñoz Sandoval, Michalina Mazurczyk, Yrene Themistocleous, Thomas A. Rawlinson, Alison Kemp, Sarah E. Silk, Jordan R. Barrett, Nick J. Edwards, Alasdair Ivens, Julian C. Rayner, Angela M. Minassian, Giorgio Napolitani, Simon J. Draper, Philip J. Spence

## Abstract

The biology of *Plasmodium vivax* is markedly different to that of *P. falciparum*; how this shapes the immune response to infection remains unclear. To address this shortfall, we inoculated human volunteers with a clonal field isolate of *P. vivax* and tracked their response through infection and convalescence. High dimensional protein and RNA-seq data show that *P. vivax* triggers an acute phase response that shares remarkable overlap with that of *P. falciparum*, suggesting a hardwired emergency myeloid response that does not discriminate parasite species. We then used cytometry by time of flight to analyse the fate and function of innate-like and adaptive T cells; these data show that *P. vivax* can activate up to one quarter of the entire T cell compartment. Heterogeneous effector memory-like CD4^+^ T cells dominate this extraordinary response and phenotypic analysis reveals unexpected features of terminal differentiation that are normally associated with cytotoxicity and autoinflammatory disease. In line with this observation, we found that CD4^+^ T cell activation coincides with collateral tissue damage and liver injury. Finally, comparative analyses demonstrate that *P. falciparum* drives T cell activation far in excess of *P. vivax*, which may partially explain why falciparum malaria more frequently causes severe disease.

## Introduction

*Plasmodium vivax* causes more than half of all malaria cases in the Americas and South-East Asia; globally, around 14 million annual cases present a significant clinical and economic burden (1). Whilst efficacious drugs are available to clear the blood-stage of infection the dormant liver stage of *P. vivax* (the hypnozoite) can persist and cause multiple relapses over many months and years. All available hypnozoitocidal drugs can cause haemolysis in patients with glucose-6-phosphate dehydrogenase (G6PD) deficiency, a highly prevalent polymorphism in many endemic populations (2, 3). This poses a unique challenge for the elimination of *P. vivax*, and forces public health services to balance the risk of potentially severe adverse treatment events (4) with the benefit of reducing malaria burden (3, 5). A vaccine that blocks parasite transmission or reduces the frequency or severity of clinical episodes would be an important step towards control and elimination, and yet vaccine development for *P. vivax* lags behind the far better studied *P. falciparum* (6). What’s more, there are large gaps in understanding the immune response to *P. vivax* and the consequences for pathology versus protection; this inevitably leads to assumptions based on the findings from falciparum malaria rather than direct experimental evidence. However, the distinct biology, pathology and epidemiology of *P. vivax* suggests that these parasites do not interact with the human immune system in the same way as *P. falciparum*. Consequently, the immune response could have an important role in shaping the discrete outcomes of vivax and falciparum malaria.

*Plasmodium vivax* is evolutionarily divergent from *Plasmodium falciparum* (7) and its genome is significantly less AT-rich. CpG islands are therefore more prevalent in *P. vivax* (8) and these DNA motifs can trigger TLR9 signalling and inflammation (9), which may partially explain why it has a much-reduced pyrogenic threshold compared to *P. falciparum* (10). Furthermore, *P. vivax* preferentially invades CD71^+^ reticulocytes (11, 12), which express class I MHC and high levels of CD47 in contrast to mature red cells (the target cell for *P. falciparum*). This provides a unique route to pathogen control in vivax malaria - the direct cytolysis of infected reticulocytes by antigen-specific CD8^+^ T cells (13). A hallmark of *P. falciparum* infected red cells is their increased rigidity, which makes them vulnerable to mechanical trapping and clearance in the spleen; in contrast, *P. vivax* infected reticulocytes become more deformable allowing for rapid egress from this site of immune surveillance (14). Importantly, *P. vivax* shows reduced cytoadherance to the endothelium and therefore sequesters less effectively than *P. falciparum*. Tissue distribution patterns are also distinct with *P. vivax* more commonly accumulating in high numbers in the bone marrow (15) and lung (16). But perhaps the most significant difference is that severe disease is a much more common outcome of falciparum malaria (17) and clinical immunity (leading to asymptomatic infection) is acquired much more quickly with *P. vivax* (18, 19). This observation was also made during the malariotherapy era, which demonstrated under controlled settings that a single episode of vivax malaria was sufficient to reduce the incidence of fever in subsequent infections (20-22).

The myriad ways in which *P. vivax* differs from *P. falciparum* therefore necessitates experimental systems and tools that are tailored specifically towards vivax malaria. This is made harder by the absence of long-term culture-adapted parasites and a consequence of this shortfall has been a resurgence in the use of human challenge models (23). Controlled human malaria infection (CHMI) was re-established for *P. falciparum* in 1986 but only very recently for *P. vivax* (24-27). To harness the exciting potential of this development we generated a new cryopreserved stabilate of *Plasmodium vivax* suitable for CHMI and used a hybrid PacBio/Illumina assembly technique to generate a reference quality parasite genome (see the accompanying article by Minassian *et al*.). Here we use systems immunology tools (including cytometry by time of flight (CyTOF)) to map the host response to *P. vivax* at unprecedented resolution. In the first instance, we tracked the acute phase response to ask how blood-stage parasites activate the innate immune system; we then examined the relationship between systemic inflammation and T cell activation. Human T cells are uniquely placed to influence the outcome of infection as they enter inflamed tissues, regulate myeloid cell activation, promote humoral immunity and even kill *Plasmodium* infected red cells (13, 28). What’s more, T cells can directly cause collateral tissue damage and pathology (29-31). Mapping the heterogeneity of T cell activation and differentiation was therefore a priority for this study. Finally, we reasoned that a comparison between the host response to *P. vivax* and *P. falciparum* may shed new light on why falciparum malaria more frequently causes severe disease and why *Plasmodium vivax* is better able to induce rapid clinical immunity.

## Results

### Plasmodium vivax triggers interferon-stimulated inflammation

Six volunteers were recruited into a clinical trial (VAC069A) to assess the infectivity of a new cryopreserved *P. vivax* stabilate, which we have called PvW1 (supplementary file 1). Importantly, these parasites originated from a naturally-infected donor in Thailand with a single genotype infection and were mosquito-transmitted prior to cryopreservation (see Minassian *et al*. for details). Each volunteer was successfully infected by direct blood challenge and reached the parasitaemic threshold necessitating treatment (5,000 or 10,000 parasite genomes ml^-1^ in the presence or absence of symptoms, respectively) within 12-16 days of inoculation (figure 1A). Whole blood samples were taken at baseline (day before challenge), during infection (e.g. C7 for 7-days post-challenge), immediately before treatment (termed diagnosis), soon after treatment (e.g. T6 for 6-days post-treatment) and 45-days after challenge (memory phase).

**Figure 1.**
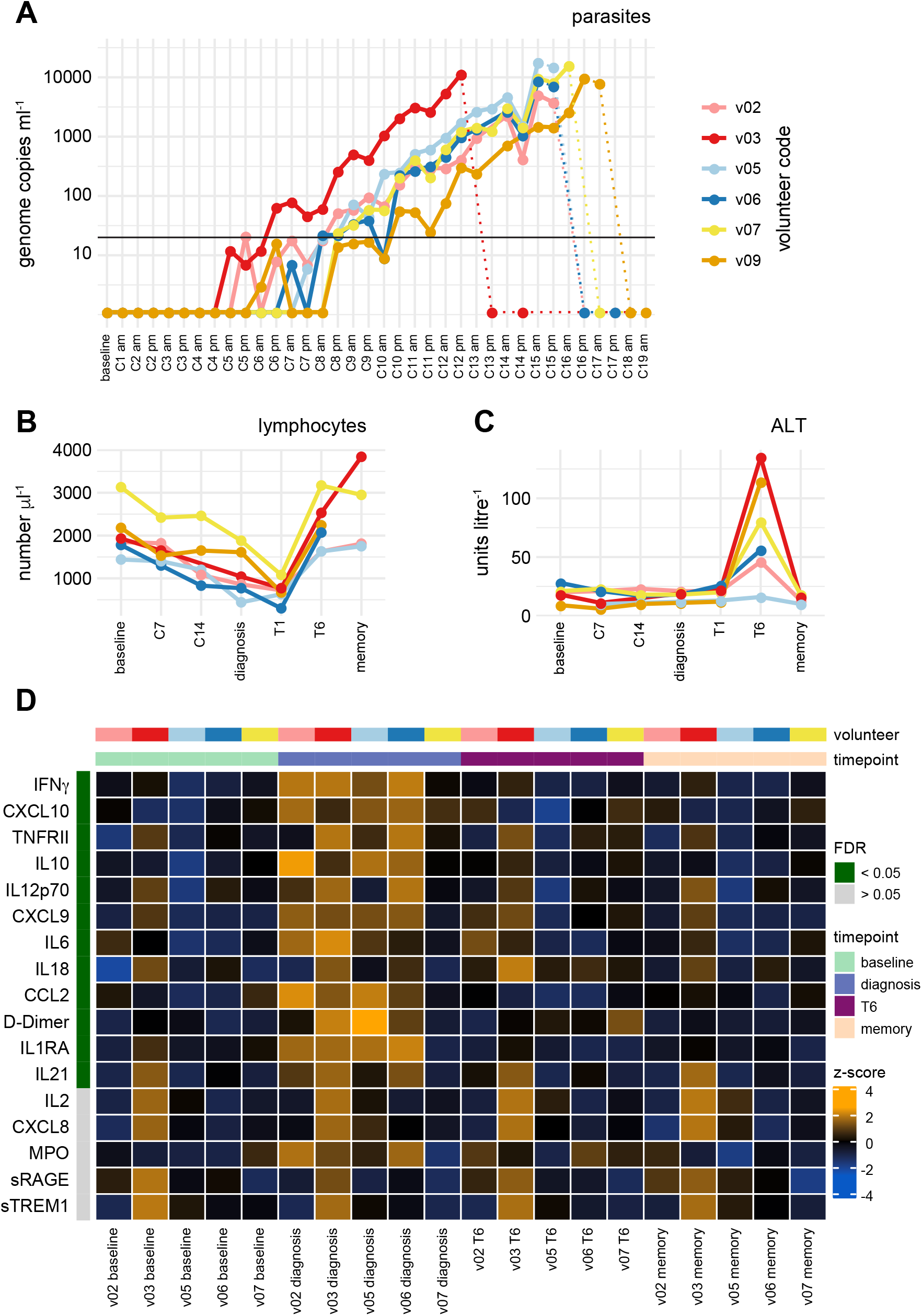
*Plasmodium vivax* triggers interferon-stimulated inflammation. (**A**) Parasitaemia was determined twice daily by qPCR. Pre-treatment measurements are shown as solid lines, post-treatment measurements as dotted lines. The limit of quantification (20 genome copies ml^-1^) is shown by a black line. (**B**-**C**) Full blood counts and blood chemistry measured (B) lymphocyte frequencies and (C) the concentration of alanine aminotransferase (ALT) throughout infection and convalescence. (**D**) Multiplexed plasma analytes were measured using a custom Legendplex assay. Each row in the heatmap is an analyte and each column a plasma sample. Samples from v09 were excluded after failing QC. Linear modelling was used to identify analytes that varied significantly through time across the cohort, and these are ordered by FDR. Only 17 of the 39 analytes measured are shown in the heatmap (those with the lowest FDR); the colour of each tile corresponds to the row-wise z-score transformed concentrations. In (B and C) the memory time-point is 90-days post-challenge and in (D) memory is 45-days post-challenge.

The total number of recorded adverse events (including symptomatology, haematology and blood chemistry) peaked within 24-hours of treatment and largely resolved by T6. All volunteers exhibited pronounced lymphopenia (an adverse event in 5/6 volunteers) and pyrexia (4/6) and thrombocytopenia (3/6) were also common features of infection (figure 1B and Minassian *et al*.). All adverse events were self-limiting and did not require specific interventions. Notably, alanine aminotransferase (ALT) increased in plasma six days post-treatment and was an adverse event in 4/6 volunteers (figure 1C). This can be indicative of hepatocellular death and liver injury - a reversible phenomenon in uncomplicated malaria (32-34) - and occurred at the same time that lymphocyte counts returned to baseline. Taken together, these results show that volunteers infected with *Plasmodium vivax* (clone PvW1) develop hallmark signs and symptoms of clinical malaria.

Next, we sought to capture the acute phase response to *P. vivax* by quantifying 39 plasma analytes including cytokines, chemokines and biomarkers of coagulation and oxidative stress using a custom bead-based array. Platelet-free plasma samples collected at baseline, diagnosis, T6 and 45-days post-challenge were used to create a time-course of abundance for each analyte. To identify analytes that varied significantly through time we fitted linear models for each analyte in the form of analyte∼timepoint+volunteer using log10 transformed concentrations and time-point as a categorical variable. After correcting for multiple testing, we found 12 analytes with an FDR < 0.05 (figure 1D).

All significant analytes were found to increase in abundance and all peaked at diagnosis with the exception of IL-18, which was significantly elevated at T6. The analyte with the lowest FDR was IFNγ indicating a type I inflammatory response, which has been extensively described in falciparum malaria (35). Key functional consequences of this response include recruitment of monocytes (CCL2) and T cells (CXCL9, CXCL10), as well as their activation and differentiation (IL-12, IL-18, IL-21). Inflammation is intimately linked to coagulation and D-Dimer (a fibrin degradation product indicative of intravascular fibrinolysis) was found to be significantly increased in all volunteers. This is a known feature of clinical malaria (36) that has been linked to disease severity (37, 38). In conclusion, *Plasmodium vivax* triggers an interferon-induced type I inflammatory response that coincides with hallmark features of clinical disease, including lymphopenia, pyrexia and fibrinolysis. Importantly, clinical presentation and plasma analytes largely return to baseline levels within six days of drug treatment with the exception of IL-18 and ALT, which are induced post-treatment and peak at T6.

### Interferon signalling is followed by a signature of cell proliferation in whole blood

To further characterise the systemic response to *P. vivax* we used bulk RNA-sequencing to resolve changes in whole blood gene expression through time. Samples were collected at baseline, throughout infection, diagnosis and T6 giving a time-course of 7-9 samples per volunteer. DESeq2 (39) was then used for pairwise comparisons to resolve conserved responses between volunteers; samples were thus grouped by time-point and compared to baseline. Differential gene expression was defined as an adjusted p value < 0.05 and an absolute fold change > 1.5 - the transcriptional response peaked at diagnosis with 2221 differentially expressed genes (DEG). Of note, no DEG were found as late as C12, which implies that the transcriptional response at diagnosis was rapidly induced. This may indicate that parasite density (rather than time elapsed) is the primary trigger for systemic inflammation in a naive host. Surprisingly, only 298 DEG were found at T6 and most of these genes (66%) were unique to this time-point, which shows that a distinct transcriptional response follows drug treatment.

To infer biological function of the host response at diagnosis and T6 we used gene ontology (GO) network analysis using ClueGO (40, 41). First, all DEG from diagnosis and T6 were combined to identify significantly enriched GO terms. Then a graph-based non-redundant network was constructed with each GO term placed as a node and with edges connecting nodes that shared associated genes. Finally, GO terms were placed into groups that have significant inter-connectivity, meaning a highly related biological function of each node within a group. Critically, for each GO term information on what fraction of associated genes are differentially expressed at diagnosis or T6 is retained; this allows for a direct comparison of the transcriptional response between time-points at a functional level. This analysis revealed diverging and disconnected biological functions at diagnosis and T6 (figure 2).

**Figure 2.**
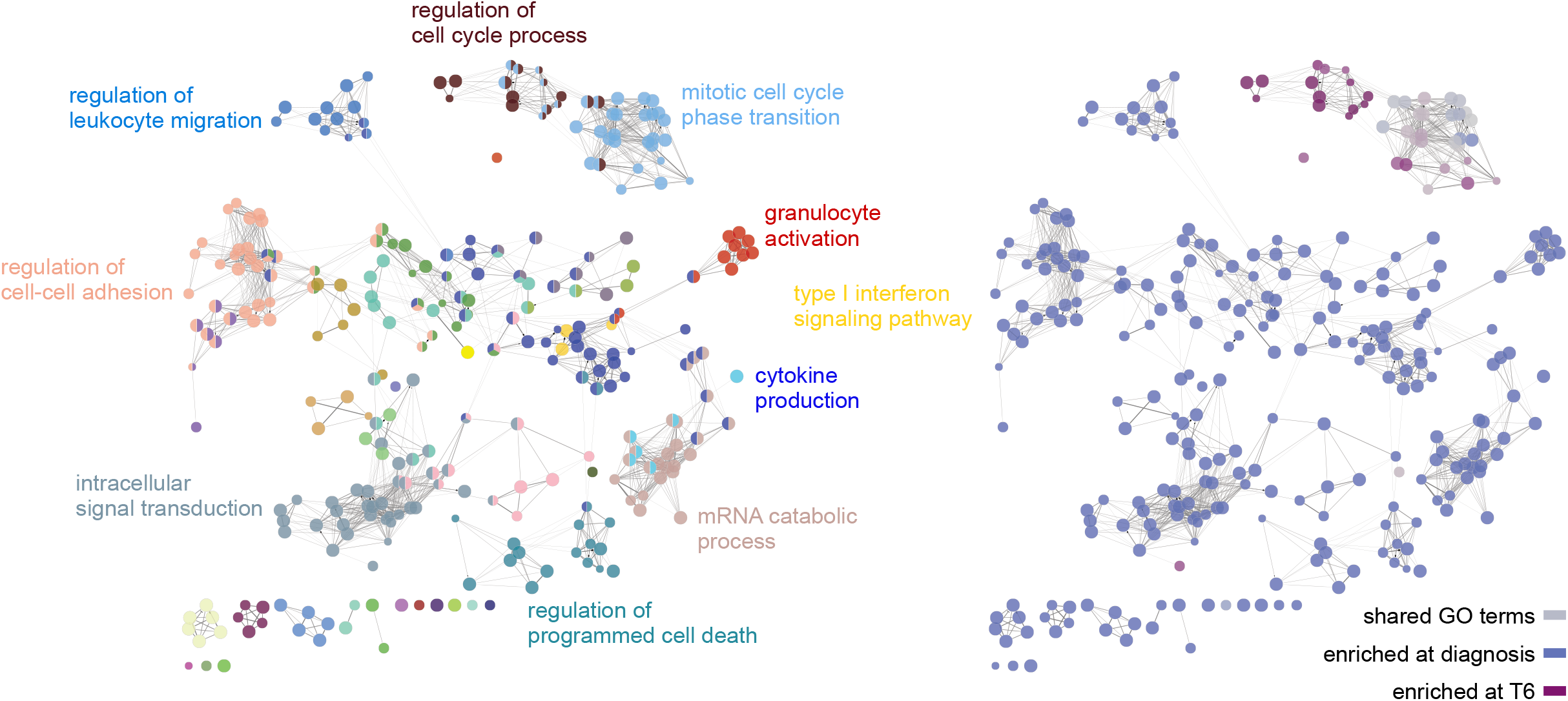
Interferon signalling is followed by a signature of cell proliferation in whole blood. Differentially expressed genes at diagnosis and T6 were combined for GO analysis. GO terms (nodes) that share associated genes are connected by an edge and edges pull GO terms into groups according to interconnectivity and overlapping function. In the left panel, each node is coloured according to functional group (named by choosing a representative GO term for each group). In the right panel, the same network is now coloured according to whether a clear majority (> 65%) of associated genes were differentially expressed at diagnosis (blue) or T6 (purple).

The transcriptional response at diagnosis is dominated by upregulation of innate signalling and defence pathways, including GO terms associated with NF-κB signalling, activation of myeloid cells, leukocyte migration and cytokine production. These pathways and functional groups were also identified when we and others analysed the transcriptional response to *P. falciparum* in naive hosts (35, 42, 43) and this signature is consistent with interferon-stimulated systemic inflammation. As volunteers are lymphopenic at this time-point the majority of these transcriptional changes are likely to derive from myeloid cells, which are initially activated by malaria parasites in the bone marrow (44). The changes we capture in whole blood during infection may therefore reveal the trafficking of activated monocytes and neutrophils as they transit from the bone marrow to inflamed tissues, such as the spleen. On the other hand, the GO terms unique to T6 all relate to cell cycle progression and nuclear division, rather than inflammation. This signature is unlikely to derive from activated myeloid cells, as in general they are terminally differentiated and do not proliferate in the circulation. Instead, these data suggest that we are capturing activated lymphocytes as they return to the circulation after parasite clearance.

In summary, *P. vivax* induces two distinct transcriptional programmes in whole blood during and after infection. During infection, transcriptional profiling reveals the rapid mobilisation of an emergency myeloid response, consistent with observations in falciparum malaria. Six days after treatment, this innate response subsides and a transcriptional signature of proliferation is revealed, which indicates widespread activation of lymphocytes.

### *Plasmodium vivax* triggers global activation of innate-like and adaptive T cells

It is well known that T cells are recruited out of the circulation during infection and activated within the inflamed spleen (45); by studying their phenotype as they re-enter the circulation at T6 it may be possible to measure T cell priming and the tissue environment in human malaria. To explore this idea further we leveraged cytometry by time of flight (CyTOF) to achieve single cell resolution of T cell activation and differentiation. Whole blood samples were preserved within 30-minutes of blood draw to create a detailed time-series of the T cell compartment and data from all volunteers were concatenated to identify conserved changes through time. To examine these data we first used Uniform Manifold Approximation and Projection (UMAP (46)) to visualise the phenotypic diversity of T cells at each time-point. Cells close to each other in the UMAP space are phenotypically similar, whereas dissimilar cells are far apart. Remarkably, the global structure of the T cell compartment appears stable between baseline and diagnosis (figure 3A) despite pronounced lymphopenia as infection progresses. At T6 however, a dense population of T cells appears *de novo* and inspection of key markers associated with lineage and memory (supplementary figure 1) indicates that these are predominantly CD4^+^ T cells with an effector memory (CD45RA^-^ CD45RO^+^CCR7^-^) and activated phenotype (CD38^hi^Bcl2^lo^) (figure 3B-C). Using the latter marker combination to identify activated T cells across all lineages, we found a surprisingly large fraction of the T cell compartment activated at T6 (up to 22% of all T cells) including innate-like gamma delta T cells and MAIT cells (figure 3D-E).

**Figure 3.**
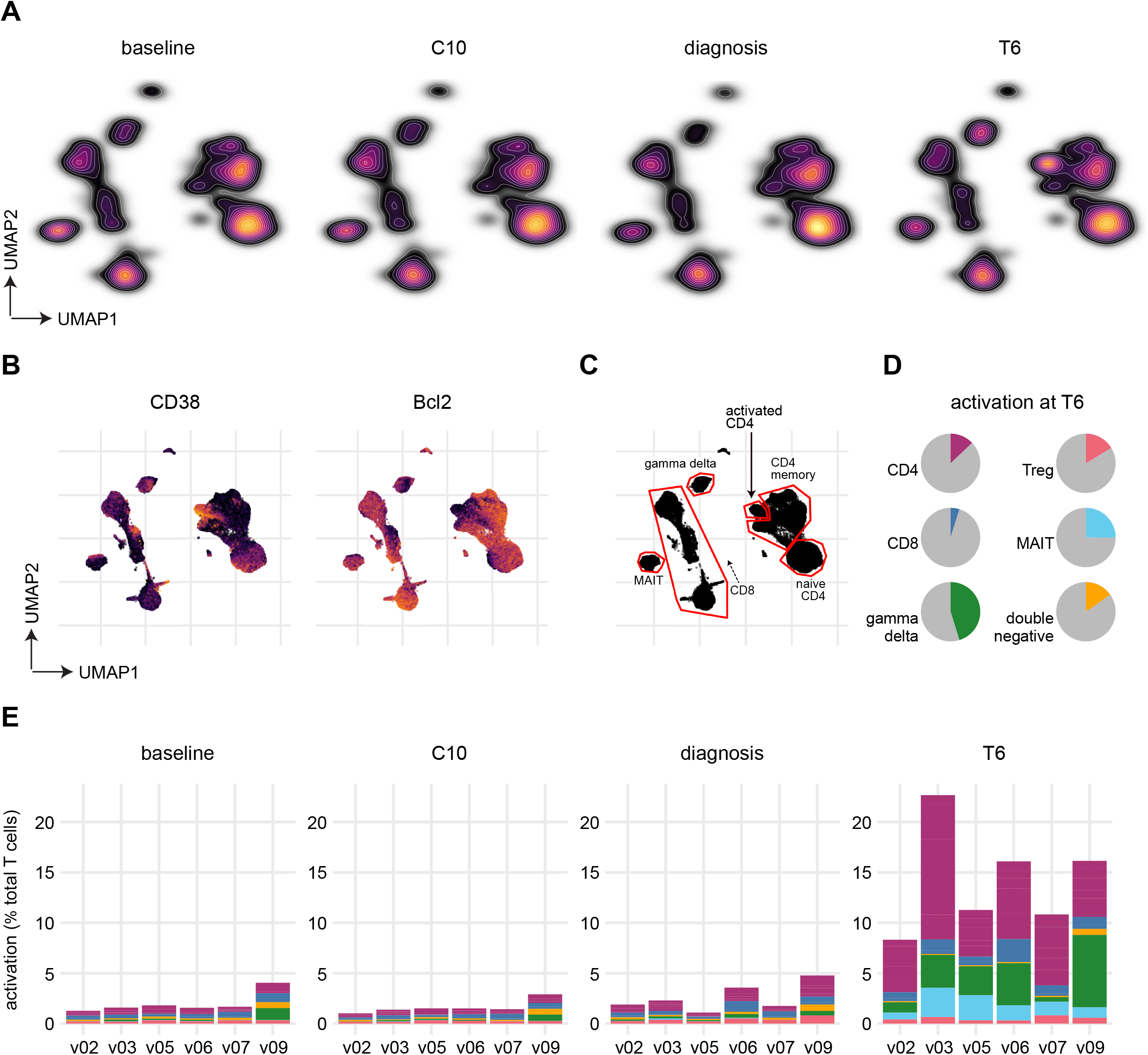
*Plasmodium vivax* triggers global activation of innate-like and adaptive T cells. Whole blood was preserved within 30-minutes of blood draw at baseline, C10, diagnosis and T6. Samples were stained with a T cell focussed antibody panel (details in supplementary file 2) and acquired on a Helios mass cytometer. (**A**) UMAP projection coloured by cell density and split by time-point. (**B**) Expression of CD38 and Bcl2 across the UMAP projection at T6; each marker is independently scaled using arcsine transformed signal intensity. (**C**) UMAP projection with gates and labels indicating the location of each major T cell subset (shown at T6, see supplementary file 1 for the expression of lineage and memory markers). Note that the dashed arrow indicates the direction of terminal differentiation for CD8^+^ T cells (e.g. TEMRA are adjacent to gamma delta T cells) (**D**) Mean proportion of each T cell subset that is activated (CD38^hi^ Bcl2^lo^) at T6. (**E**) Stacked bar chart showing the frequency of activated T cells at each time-point; bars are colour-coded by lineage as in (D).

In order to more comprehensively describe phenotypic changes in T cells we used FlowSOM clustering (47) and manual merging to assign each T cell to one of 34 non-overlapping clusters (supplementary figure 2). Tracking the frequency of each cluster through time then resolved the dynamic changes in the T cell compartment as a consequence of infection (supplementary figure 3). To statistically determine which clusters vary significantly through time we performed linear regression on cell count data using *edgeR* (48, 49). In this way, we could identify differentially abundant clusters at each time-point relative to baseline. We found no clusters differentially abundant at C10 and only one cluster (CD161^+^ gamma delta) downregulated at diagnosis (FDR < 0.05 and absolute fold change > 2). That only one of 34 clusters significantly changed in their relative size as the host became lymphopenic indicates that T cells are proportionally recruited out of the circulation regardless of lineage, function or memory, suggesting a relatively indiscriminate mechanism of recruitment.

Using the same significance cut-offs we identified nine clusters that increased in abundance at T6 (figure 4A-B) - these clusters were comprised of five CD4^+^ and two CD8^+^ T cell subsets, plus one MAIT and one gamma delta subset. Critically, all displayed a CD38^hi^Bcl2^lo^ phenotype (figure 3B and figure 4A) statistically confirming widespread activation of all T lineages. Notably, although a cluster of activated regulatory T cells increased in abundance at T6 this did not reach statistical significance, most likely due to our small sample size. Collectively, these results suggest that innate-like and adaptive T cells are indiscriminately recruited out of circulation and activated by *P. vivax*. The breadth and scale of T cell activation considerably exceeds what has been observed in other human challenge models, including typhoidal *Salmonella* (50) and influenza A (51).

**Figure 4.**
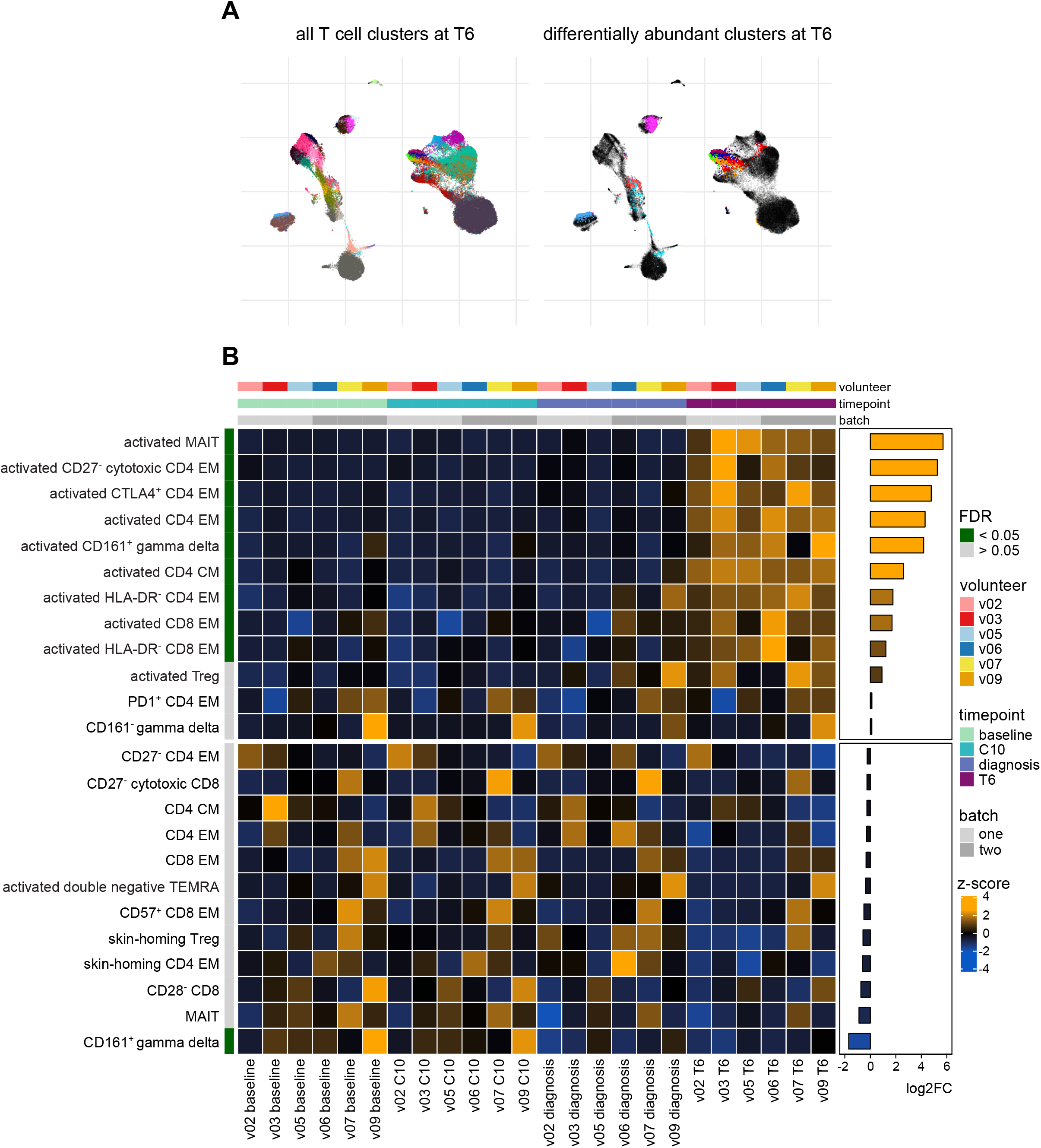
Differential abundance of T cell clusters through time. (**A**) UMAP projection showing all 34 T cell clusters (left) and those that were differentially abundant at T6 (right). Each cluster has a unique colour. (**B**) Heatmap showing the relative abundance of T cell clusters through time. Each row is a T cell cluster and each column a sample; clusters are ordered by log2 fold change at T6 (relative to baseline). Only 24 of the 34 T cell clusters are shown (those with the lowest FDR) and tiles are coloured according to row-wise (arcsine square root transformed) frequency z-scores.

### Heterogeneous effector memory-like CD4^+^ T cells dominate the adaptive response

In order to elucidate the function of activated T cells as they re-enter the circulation we inspected the median expression values of proliferation and differentiation markers in the nine T cell clusters that increased in abundance at T6 (figure 5A and supplementary figure 4). And because we found more than half of all activated T cells were CD4^+^ (with five distinct clusters contained within this lineage (figure 5B)) we examined in detail the heterogeneity of the CD4^+^ T cell response. High CD38 expression and low Bcl2 expression was a shared feature of all significant clusters and 4/5 displayed an effector memory phenotype (CD45RA^-^ CD45RO^+^CCR7^-^). The one exception was a small cluster of activated central memory-like CD4^+^ T cells (CD45RA^-^CD45RO^+^CCR7^+^). By summing these five CD45RO^+^ clusters we found that *P. vivax* activates 20-35% of the non-naive CD4^+^ T cell pool (figure 5C).

**Figure 5.**
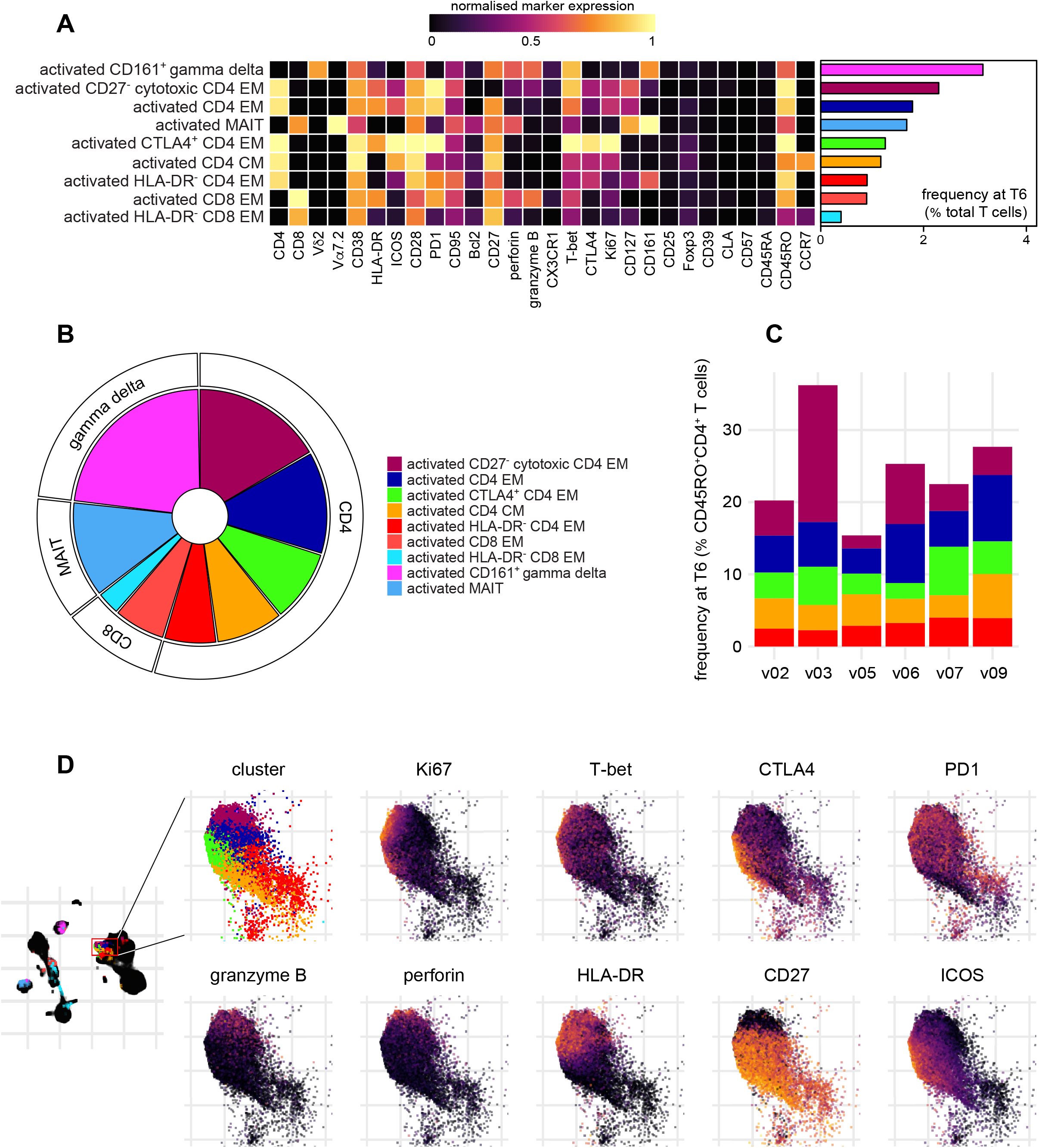
Heterogeneous effector memory-like CD4^+^ T cells dominate the adaptive response. (**A**) Heatmap showing normalised median expression values of all markers used for clustering in each of the 9 T cell clusters that were differentially abundant at T6; the horizontal bar chart shows the average frequency of each cluster across all volunteers. (**B**) Pie showing the relative size of each differentially abundant T cell cluster.(**C**) Stacked bar chart showing each differentially abundant CD4^+^ T cell cluster as a proportion of the total non-naive (CD45RO^+^) CD4^+^ T cell pool. (**D**) UMAP projection showing the expression of activation, proliferation and differentiation markers across each of the differentially abundant CD4^+^ T cell clusters at T6; each marker is independently scaled using arcsine transformed signal intensity.

The T cell activation markers HLA-DR and ICOS were a common (but not ubiquitous) feature of activated CD4^+^ T cells and we also found widespread expression of inhibitory receptors, such as PD1 and CTLA4 (figure 5D). These checkpoint inhibitors are often used to identify exhausted T cells and yet our volunteers experienced a relatively short and acute infection (rather than chronic stimulation). However, it is now recognised that upregulation of these receptors can also act to self-regulate short-lived responses (52, 53). In agreement, we found that many activated CD4^+^ T cells were CD28^hi^, T-bet^+^ and proliferative (Ki-67^+^) indicating polarisation towards a functional and inflammatory TH1 fate (figure 5A).

Perhaps surprisingly, the largest cluster of activated CD4^+^ T cells had a CD27^-^ cytotoxic phenotype (perforin^+^ granzyme B^+^) (figure 5A, B and D). Loss of CD27 expression and gain of cytotoxic functions is not a common outcome of CD4^+^ T cell activation in human infectious disease. Rather, this phenotype usually arises in contexts of persistent antigen stimulation (54) or in conditions associated with collateral tissue damage, such as pulmonary tuberculosis (55), IgG4-related disease (56) or rheumatoid arthritis (57). Whether this phenotype is specific to *Plasmodium vivax* or is a shared feature with *P. falciparum* remains to be seen. In summary, CD4^+^ T cells with an effector memory phenotype dominate the adaptive response to *P. vivax* and display marked heterogeneity in their expression of key markers of T cell function and fate (figure 5D). These data therefore emphasise the complexity of CD4^+^ T cell activation and differentiation in vivax malaria.

### T cell activation occurs independently of systemic inflammation

Innate-like and adaptive T cells have distinct ligand requirements for TCR signalling and yet every major T cell subset is activated by *P. vivax* (figure 4A-B). We therefore hypothesised that the scale and breadth of the T cell response may indicate bystander (antigen-independent) activation, which can be caused by systemic inflammation (58-61). To investigate the relationship between the interferon-induced type I inflammatory response observed at diagnosis and T cell activation at T6 we constructed a Pearson correlation matrix (figure 6A). We input the log2 fold change of each plasma analyte with an FDR < 0.05 and the log2 fold change of each activated T cell cluster (defined as CD38^hi^Bcl2^lo^). To examine all possible correlations we chose to include all activated T cell clusters regardless of whether their abundance was significantly increased or not. Fold change was calculated for each feature at either diagnosis or T6 (relative to baseline) depending on when the peak response was observed. Hierarchical clustering on the matrix revealed extensive positive correlation between inflammatory cytokines, chemokines and coagulation at diagnosis. In contrast, only one T cell cluster correlated highly with these analytes (r > 0.8 for activated CD8 effector memory) and just two clusters showed weak correlations (activated CD161^+^ gamma delta and activated CD4 effector memory). Instead, the majority of T cell clusters (8/11) were placed into a separate clade together with ALT, which indicates that the majority of the T cell response is co-regulated but operates independently of systemic inflammation.

**Figure 6.**
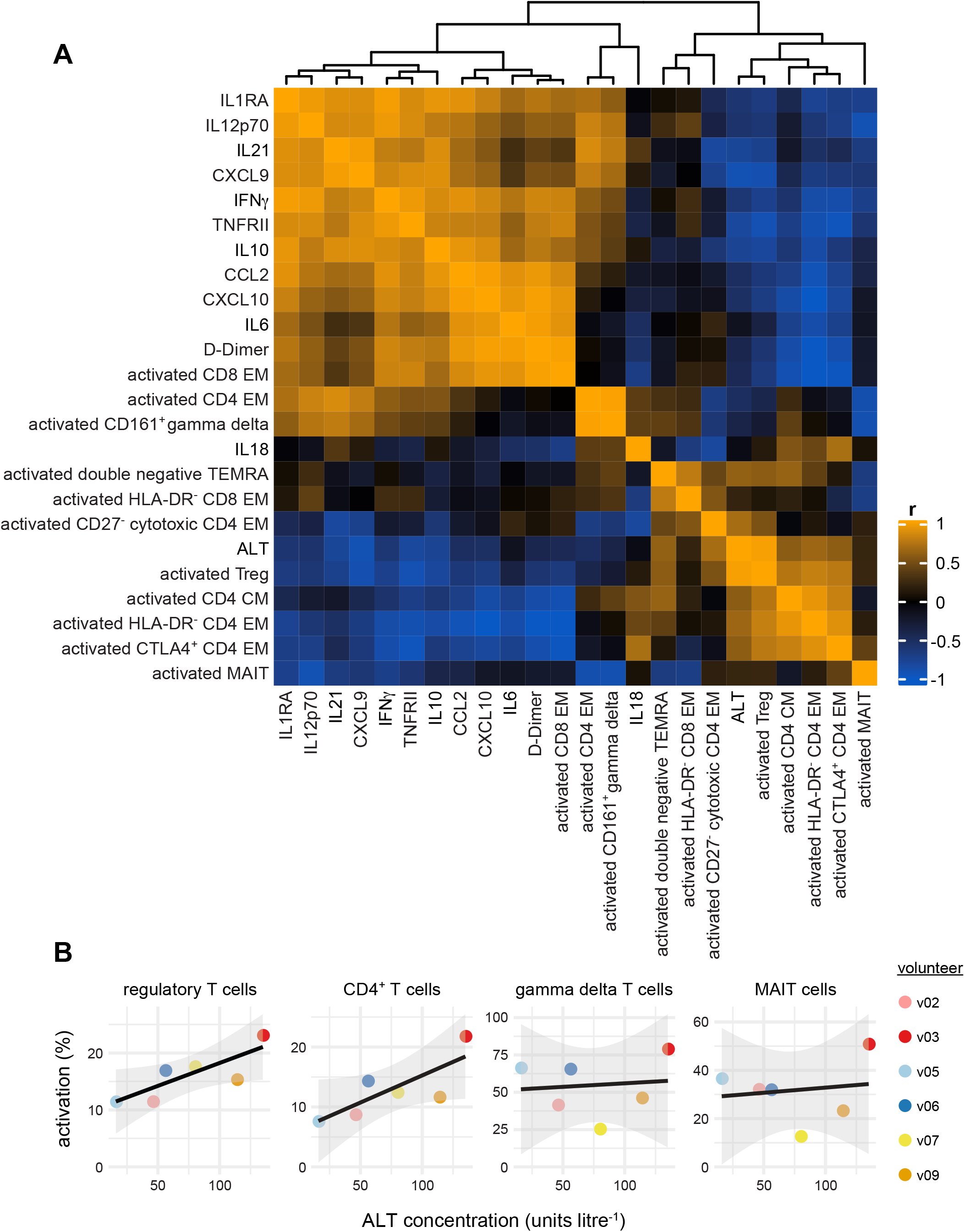
T cell activation occurs independently of systemic inflammation. (**A**) Heatmap showing a Pearson correlation matrix of the log2 transformed fold change of each activated T cell cluster and the twelve most variable plasma analytes (FDR < 0.05). Fold change was calculated either at diagnosis or T6 (relative to baseline) according to when this was largest for each feature. The absolute concentration of plasma ALT at T6 (the peak of the response) is also included. The order of features was determined by hierarchical clustering and the associated dendrogram is shown at the top of the heatmap. (**B**) Correlation between ALT concentration and the activation of innate-like and adaptive T cells at T6. Loess regression line is shown in black and the 95% confidence intervals in grey.

We next looked at the co-regulation of T cell activation and ALT in more detail. Elevations in plasma ALT showed a highly positive correlation with expansion of activated Tregs (r = 0.97) and a modest positive correlation with four activated CD4^+^ T cell clusters, indicating a possible association between T cell activation and collateral tissue damage. Because this analysis was looking for independent relationships between each T cell cluster and ALT we decided to repeat this analysis at a subset level. To this end, we calculated the correlation between lineage-specific T cell activation and absolute ALT levels at T6 and found a significant or near significant association with activated CD4^+^ T cells (r = 0.791, p = 0.061) and regulatory T cells (r = 0.816, p=0.0478) but not innate-like MAIT (r = 0.147, p= 0.781) or gamma delta T cells (r = 0.107, p = 0.841) (figure 6B). In summary, there is no clear relationship between the intensity of systemic inflammation at diagnosis and the magnitude of the T cell response at T6. In contrast, activation of CD4^+^ and regulatory T cells positively correlates with elevations in circulating ALT, which suggests that the adaptive T cell response to *P. vivax* may be an accurate indicator of collateral tissue damage.

### Parasite species regulates CD4^+^ T cell activation

Last of all, we wanted to investigate whether the immune response to *P. vivax* differed from that to the far better studied pathogen *P. falciparum*. Initially, we compared transcriptional signatures in whole blood between volunteers infected with *P. vivax* (this study) and *P. falciparum* (a CHMI trial we conducted previously with time-matched samples (VAC063C)). Importantly, volunteers in both studies were malaria-naive, infected by blood challenge and diagnosed and drug treated at the same parasite density (see methods). Differentially expressed genes (adj p < 0.05 and absolute fold change > 1.5) were identified at diagnosis and T6 (relative to baseline) in both volunteer cohorts using DESeq2. The DEG from each cohort were then combined at each time-point for GO term enrichment. Crucially, information was retained to indicate what fraction of associated genes for each GO term derived from *P. vivax* or *P. falciparum* infected volunteers. At diagnosis we found 289 GO terms of which 282 (97.58%) were shared almost equally between cohorts (figure 7A). ClueGO network construction then revealed that these shared GO terms organised into functional groups related to myeloid cell activation and systemic inflammation (figure 7B). Remarkably, we found only 7 GO terms (2.42%) with associated genes that majoritively (> 65%) derived from one volunteer cohort. All of these cohort-specific GO terms were located in the same region of the ClueGO network and were enriched in volunteers infected with *P. vivax* - this response was characterised by downregulation of structural ribosomal gene expression, which can be induced by type I interferon signalling (62). Despite this difference, these results show that the functional consequences of the innate immune response are highly conserved between two divergent parasite species, which differ in host cell tropism, red cell remodelling, tissue sequestration and in their potential to cause severe disease.

**Figure 7.**
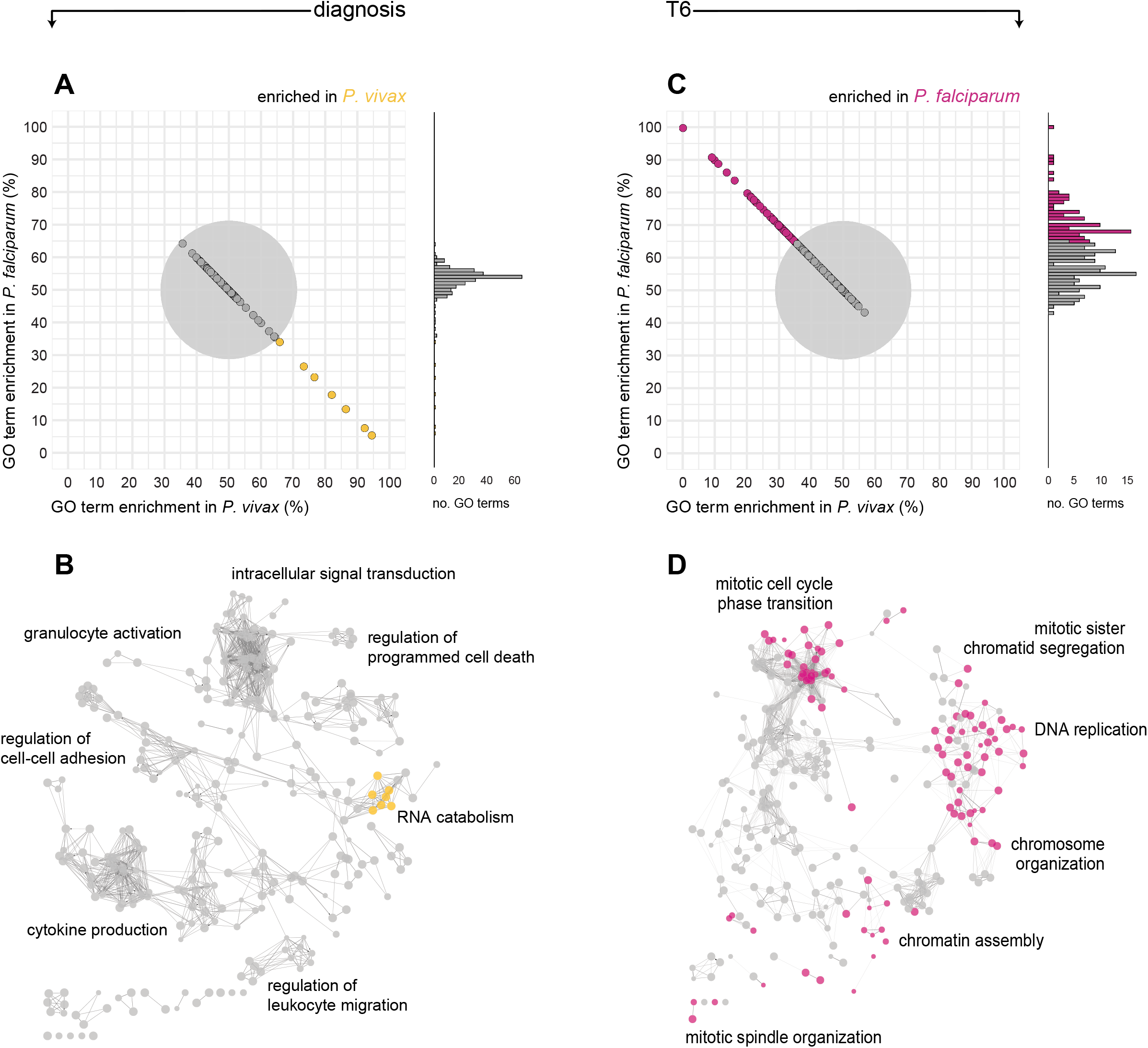
The innate response to malaria does not distinguish parasite species. Whole blood RNA-sequencing was performed analogously during the VAC069A (*P. vivax*) and VAC063C (*P. falciparum*) CHMI studies (n = 6 and 3, respectively). Differentially expressed genes in *P. vivax* and *P. falciparum* were combined for GO analysis at diagnosis (A and B) or T6 (C and D). (**A** and **C**) Each GO term is represented by a point whose position on the x and y axis is determined by what proportion of associated genes were differentially expressed in volunteers infected with *P. vivax* or *P. falciparum*, respectively. The grey circle represents a 65% threshold that needed to be crossed to call a GO term as majoritively associated with one volunteer cohort. Because many points overlap around 50% a histogram of the number of GO terms is also shown along the y axis. All points and bars are coloured according to enrichment. (**B** and **D**) ClueGO networks of the GO terms represented in (A and C); nodes are coloured according to enrichment in *P. vivax* or *P. falciparum*. For each of the major functional groups a representative GO term was chosen as group name.

To examine parasite species-specific differences in the adaptive immune response to malaria we repeated this analysis at T6. In contrast to diagnosis, only 151/235 (64.3%) GO terms were shared between volunteer cohorts at this time-point (figure 7C). These shared features related to nuclear activities involved in cell division, such as sister chromatid segregation and mitotic spindle organization (figure 7D). On the other hand, 84/235 (35.7%) GO terms were predominantly derived from just one dataset (*P. falciparum*) and these terms were accessory to cell division, such as DNA replication and mitotic cell cycle phase transition. It is therefore apparent that in both datasets the predominant transcriptional signature six days post-treatment is one of cell division but the transcriptional changes induced by *P. falciparum* are significantly more pronounced. This may indicate that *P. falciparum* can induce lymphocyte activation on a scale in excess even of *P. vivax*.

To explore this possibility we examined the induction of signature genes associated with T cell activation and differentiation in whole blood (figure 8A). Remarkably, we could detect the upregulation of all major hallmarks of T_H_1 polarisation (including T-bet and IFNγ) in volunteers infected with *P. falciparum* (but not *P. vivax*). We then directly compared the activation of CD4^+^ T cells between volunteers by CyTOF (figure 8B-C). Whole blood samples were collected, preserved and processed for mass cytometry in exactly the same way during the VAC069A and VAC063C trials. FlowSOM clustering was performed independently for each cohort and we summed all activated (CD38^hi^Bcl2^lo^) clusters to measure the relative size of the adaptive response. This analysis clearly showed that *P. falciparum* drives increased activation of CD4^+^ and regulatory T cells in malaria-naive hosts.

**Figure 8.**
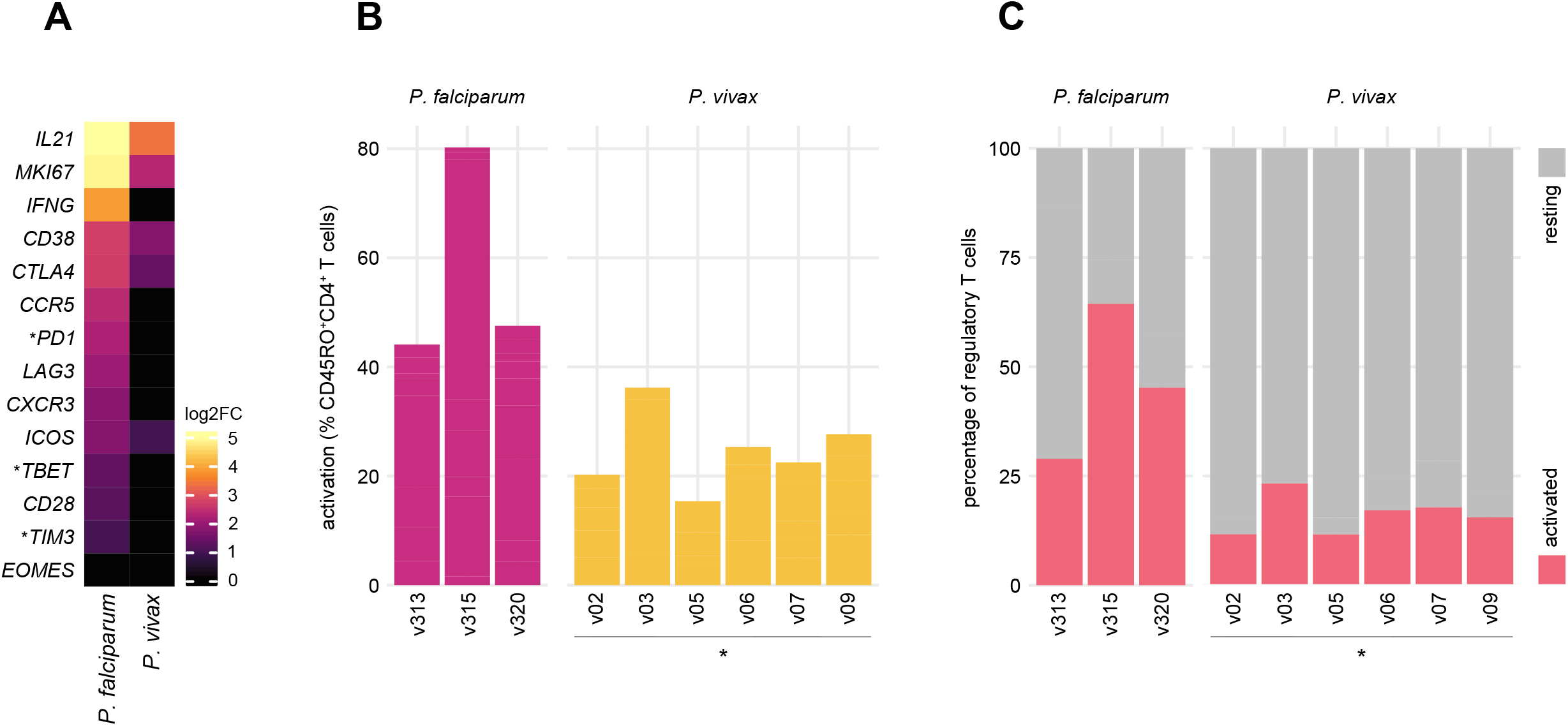
Parasite species regulates CD4^+^ T cell activation. (**A**) Heatmap of signature T cell genes showing their log2 fold change at T6 (relative to baseline) in whole blood; asterisks indicate that common gene names were used. Genes that were not differentially expressed (adj p > 0.05) are shown with a fold change of zero. (**B**) Proportion of non-naive (CD45RO^+^) CD4^+^ T cells that are activated (CD38^hi^Bcl2^lo^) at T6 in volunteers infected with either *P. falciparum* or *P. vivax* by CHMI. (**C**) Proportion of regulatory T cells that are activated at T6 in volunteers infected with *P. falciparum* or *P. vivax*. In (B and C) FlowSOM was used to identify activated T cell clusters (CD38^hi^Bcl2^lo^) independently in each volunteer cohort; the frequency of activated clusters were summed and are shown as a proportion of the indicated lineage (analogous to figure 5C). Note that all volunteers were malaria-naive and therefore experienced a first-in-life infection. * p = 0.02381 Wilcoxon rank sum exact test (two-tailed).

In summary, the acute phase response that leads to systemic inflammation and coagulation is shared between falciparum and vivax malaria, despite vast differences in parasite biology, pathology and epidemiology. This suggests that innate cells do not differentiate between parasite species and simply launch a hardwired programme of activation. In agreement, every plasma analyte significantly upregulated in naive hosts infected with *P. vivax* (figure 1D) is also upregulated during CHMI with *P. falciparum* (VAC063C, Muñoz Sandoval *et al*., in preparation). In contrast, the adaptive immune response (measured after drug treatment and the release of lymphocytes from inflamed tissues) differs markedly. Our data indicate that *P. falciparum* drives increased CD4^+^ T cell activation as compared to *P. vivax*, which is already capable of activating one quarter of the entire T cell compartment.

## Discussion

The innate immune response to *P. vivax* and *P. falciparum* appears to be indistinguishable at a functional RNA and protein level. Both responses are dominated by activation of myeloid cells, likely driven by interferon signalling, the abundant release of diverse proinflammatory molecules, leukocyte migration, coagulation and hallmark signs and symptoms of clinical malaria. Circulating monocytes and neutrophils are largely recent emigrants from the bone marrow and recent studies have shown that innate priming during the blood-stage of infection first occurs in this tissue (44, 63). *P. vivax* and *P. falciparum* differ significantly in their genome composition, host cell tropism and prevalence in the bone marrow (64, 65); that these differences have no apparent bearing on the initiation of systemic inflammation is surprising and suggests that the emergency myeloid response is hardwired. This finding is analogous to our recent report that two genetically distinct *P. chabaudi* clones, which vary markedly in their capacity to sequester and cause hyperparasitaemia and severe disease, trigger near-identical myeloid responses in the bone marrow and spleen (66).

These data have important implications for studies that attempt to resolve drivers of severe malaria. First, if there is no difference between *P. vivax* and *P. falciparum* in how they activate the innate immune system then innate responses are unlikely to be sufficient for the severe manifestations of disease, an outcome largely confined to falciparum malaria. That is not to say that innate inflammation does not promote severe disease but instead point out that excessive innate responses must be driven by additional extrinsic factors, such as pathogen load (generally higher with *P. falciparum* (67, 68)) or T cell activation. Second, studying activation of innate immune cells in the circulation (and the abundance of systemic inflammatory markers) is not a good indicator of tissue-specific responses. This is evidenced by the clear disconnect we identify between systemic inflammation and the level of T cell activation and tissue damage measured after resolution of the acute phase response. This finding could in part explain why the association between inflammatory markers measured in blood and disease outcome are often so contradictory in endemic settings (69). We therefore suggest that field studies should examine the function and fate of adaptive immune cells when they return to the circulation and this can be readily achieved by extending sampling for immune profiling beyond drug treatment.

We found innate-like and adaptive T cells express a broad range of activation markers and the marker combination CD38^hi^Bcl2^lo^ was sufficient to identify activated cells in every lineage. Cumulatively, the activated fraction of total T cells far exceeds what has been reported for other human pathogens (50, 51); this indicates that T cells are indiscriminately activated by malaria parasites. In this context, it is particularly significant that MAIT cells were activated since they do not recognise peptide antigens loaded onto MHC molecules; instead, they recognise riboflavin metabolite-derived antigens presented by APCs expressing MR1 (70). *Plasmodium spp*. lack the pathways for riboflavin synthesis, suggesting that MAIT cells are not responding in an antigen-specific manner. These innate-like cells can act as sensors of tissue inflammation and be directly activated by cytokines such as IL-12 and IL-18 (61, 71, 72). Adaptive T cells can also be activated and clonally expand via this route (59, 60, 73). Our data therefore raise the intriguing possibility that *P. vivax* triggers polyclonal bystander T cell activation in naive hosts. TCR sequencing has shown polyclonal expansion of effector CD4^+^ T cells in *Plasmodium chabaudi*, which lends support to this hypothesis (74).

CD4^+^ T cells dominated the adaptive response to *P. vivax* and all significant clusters had a memory phenotype 6-days after drug treatment. But are these *bona fide* memory cells? This may depend upon their antigen specificity; if all activated CD4^+^ T cells recognise *Plasmodium* epitopes then they may simply be short-lived effectors and the number that persist to form a memory reservoir would need to be quantified further into convalescence. On the other hand, if CD4^+^ T cells specific for other pathogens (or vaccine epitopes) are activated this might indicate clonal expansion of pre-existing memory cells (58). Compared to naive T cells, memory cells are more easily activated in the absence of TCR signals (75) - future studies should therefore prioritise investigating the antigen specificity and clonality of activated T cells in human malaria. In the meantime, our data show that (regardless of specificity) the differentiation of CD4^+^ T cells towards an inflammatory T_H_1 fate is a common outcome of infection in naive hosts; this is evident in the widespread expression of T-bet, Ki67, ICOS and CD28 in activated cells. Although most activated cells also expressed the inhibitory receptors PD1 and CTLA4 this is now recognised to be an essential mechanism of self-regulation (52, 53) - expression of PD1 cannot therefore serve as shorthand for T cell exhaustion in malaria. Surprisingly, the major activated CD4^+^ T cell cluster displayed a CD27^-^ cytotoxic phenotype; cells pushed down this route of differentiation typically arise in the context of chronic stimulation or in autoinflammatory diseases (54-57). These data therefore highlight the potency of the tissue environment for driving T cell activation in vivax malaria.

More broadly, our data highlight the heterogeneity of activated CD4^+^ T cells; clustering identified at least five distinct populations, which underlines the power of high-dimensional cytometry in uncovering cell fates that can easily be missed when relying on just a few markers to define complex phenotypes. Future studies that incorporate additional transcription factors and intracellular cytokines in the staining panel are likely to reveal even richer functional diversity. Furthermore, the results presented in this study reveal only the conserved T cell responses that are shared between volunteers. Human T cells are inherently plastic (76) and there is enormous variation in the immune response to pathogens and their products (77, 78). Immune variation is therefore likely to play a key role in determining disease outcome in human malaria (35) and future studies can address this point by increasing sample size and applying individual (rather than group-level) analysis techniques to study T cell fates.

This study had a number of additional limitations. We cannot exclude effects of drug treatment as a confounding variable at post-treatment time-points. Direct xenobiotic effects seem unlikely to drive the T cell activation we observe, however, as artemisinin and its derivatives have been shown to inhibit T cell responses in a dose-dependent manner both *in vitro* and *in vivo* (79, 80). Drug treatment is also unlikely to explain the increase in ALT observed in most volunteers. Liver injury is a common feature of clinical and experimental malaria and occurs independently of the drug used or the treatment regime (32-34). Furthermore, abnormal ALT levels significantly decrease 7-days after drug treatment in patients with severe malaria (81), suggesting that infection or inflammation is the driver of hepatocellular death. Of note, there were no significant deviations in albumin or bilirubin in our volunteer cohort (Minassian *et al*.) indicating that the biosynthetic and metabolic functions of the liver have not been compromised. Elevations in ALT are therefore a marker of asymptomatic collateral tissue damage in CHMI, as previously suggested (33). Another limitation of this study is that infections had to be terminated at a parasite density much lower than would be observed in endemic settings. It is possible (perhaps likely) that we would have found the innate immune response to *P. vivax* and *P. falciparum* diverge later in infection although this would not contradict our conclusion that innate cells have no intrinsic capacity to differentiate between parasite species. It is also unclear whether the levels of T cell activation observed here can be maintained in infections of longer duration, or how chronic infection would alter T cell function.

Our data nevertheless indicate that early in infection *P. falciparum* exceeds the already prolific *P. vivax* in triggering CD4^+^ T cell activation and proliferation independently of pathogen load. This is evidenced by the remarkable transcriptional signature of T_H_1 polarisation in whole blood when volunteers are infected with *P. falciparum* (but not *P. vivax*) despite using the same study end-points. Moreover, CD4^+^ T cell activation (including regulatory T cells) was closely associated with tissue damage (ALT). As such, if *P. falciparum* is better able to drive T cell activation and tissue damage in naive hosts then it is tempting to speculate that this may be one way in which falciparum malaria causes more severe disease (82).

Finally, all volunteers recruited to this study were malaria-naive and so some of the features we describe here could be specific to first-in-life infections. This is conceivable since both epidemiological and experimental data indicate that first infection differs from reinfection in regards to the pyrogenic threshold (22, 83) and likelihood of severe malaria (84). In this context, human rechallenge models may reveal how the immune response evolves to repeated infections and could identify adaptations that coincide with clinical immunity. This would be a powerful and complementary approach to the comparison of human T cell responses between *P. vivax* and *P. falciparum*. And together these approaches to experimental medicine may help resolve the immune mechanisms that can reduce disease burden.

## Methods

### Clinical Trial Design

Six volunteers were recruited to test the infectivity and safety of a new cryopreserved stabilate containing a clonal field isolate of *P. vivax* (PvW1); this stabilate had carefully been prepared for use in CHMI by blood challenge (Minassian *et al*.). These six volunteers were enrolled into the VAC069 study and termed cohort A; the CHMI trial itself was named VAC069A. VAC069A was sponsored by the University of Oxford, received ethical approval from UK NHS Research Ethics Service (South Central - Hampshire A reference 18/SC/0577) and was registered on ClinicalTrials.gov (NCT03797989). The trial was conducted in line with the current version of the Declaration of Helsinki 2008 and conformed with the ICH Guidelines for Good Clinical Practice.

The VAC069A trial is reported in full in the accompanying article by Minassian *et al*. In brief, cryopreserved vials of stabilate were thawed, washed and diluted under aseptic conditions, and then administered to healthy malaria-naive adult volunteers by intravenous injection. Volunteers each received an inoculum containing between 116 and 2232 PvW1 genome copies; variation in copy number had no measurable effect on parasite multiplication rate (supplementary file 1). It is important to note that parasite dose may influence the time to diagnosis but has no effect on outcome of infection in human malaria (85). After inoculation whole blood was drawn twice daily to determine parasitaemia by qPCR (target gene = 18S ribosomal RNA). Thick blood smears were also evaluated by experienced microscopists at each time-point. Treatment was initiated once two diagnostic conditions were fulfilled: parasitaemia above 5,000 parasite genome copies ml^-1^, parasitaemia above 10,000 genome copies ml^-1^, positive thick blood smear and/or symptoms consistent with malaria. Treatment usually consisted of artemether and lumefantrine (Riamet) or atovaquone and proguanil (Malarone) if Riamet was contraindicated. Volunteer 05 received Malarone, all other volunteers received Riamet.

All reported clinical symptoms (arthralgia, back pain, chills, diarrhoea, fatigue, fever, headache, malaise, myalgia, nausea, pyrexia, rigor, sweating, vomiting) were recorded as adverse events and assigned a severity score: 1 - transient or mild discomfort (no medical intervention required); 2 - mild to moderate limitation in activity (no or minimal medical intervention required); 3 - marked or severe limitation in activity requiring assistance (may require medical intervention). At baseline, C7, C14 (if undiagnosed), diagnosis, T1 and T6 full blood counts and blood chemistry were evaluated at the Churchill and John Radcliffe Hospital in Oxford, providing 5-part differential white cell counts and quantification of electrolytes, urea, creatinine, bilirubin, alanine aminotransferase (ALT), alkaline phosphatase (ALP) and albumin.

Blood for immunological analyses was collected in EDTA tubes by venepuncture at the indicated time-points. All samples were processed immediately for downstream applications in a laboratory adjacent to the clinical facility.

### Multiplexed plasma analyte analysis

Whole blood was centrifuged at 1000g for 5 minutes to separate cellular components and plasma. Plasma was then aspirated and centrifuged at 2000g for 10 minutes to remove platelets. Avoiding the pellet, supernatants were aliquoted and snap-frozen on dry ice before storage at -80°C until further processing. Plasma samples from baseline, diagnosis, T6 and 45-days post-challenge (memory phase) were thawed on ice before centrifuging at 1000g for 1 minute to remove potential protein aggregates. The concentration of 39 different analytes was then measured by running every sample across four different custom Legendplex assays from Biolegend, according to the manufacturer’s instructions. Filter plates with samples and concentration standards were then acquired on a LSRFortessa flow cytometer (BD). FCS files were processed using Legendplex software (version 7.1), which automatically interpolates a standard curve using the plate-specific standards and calculates analyte concentrations for each sample. Samples from v09 were excluded after failing QC and statistical analysis of the remaining 5 volunteers was carried out in R (version 3.6.3). Using *stats*, linear models were fit using restricted maximum likelihood for each analyte with log10 transformed analyte concentrations as response variable and time-point and volunteer as categorical fixed effects. Linear hypothesis testing via pairwise comparisons was performed using *multcomp*’s glht function and adjusted for multiple testing (Benjamini & Hochberg). An FDR < 0.05 was considered significant. Results were visualised using *ComplexHeatmap* (86), inspired by the plotDiffHeatmap function of *CATALYST* (87) and *ggplot2* (88).

### RNA-sequencing and data analysis

Whole blood was added to Tempus reagent (Applied Biosystems) at a ratio of 1:2 within 60-minutes of blood draw before storage at -80°C. RNA extraction was performed using the Tempus Spin RNA isolation reagent kit (Applied Biosystems) according to the manufacturer’s instructions. Briefly, lysed blood samples were thawed and centrifuged at 3000g for 30 minutes at 4°C to pellet nucleic acids. Pellets were resuspended in RNA purification re-suspension solution and centrifuged on a silica column to remove non-nucleic acid contaminants. After washing, the column was incubated for two minutes at 70°C before eluting nucleic acids. The eluate was then subjected to DNA digestion using the RNA Clean and Concentrator-5 kit (Zymo Research). Purified RNA was eluted in 30µl DNAse/RNAse-free water; quantification and quality control were carried out on a Qubit (Thermo Fisher Scientific) and Bioanalyzer (Agilent Technologies), respectively. All samples were diluted to a concentration of approximately 20-40 ng/µl and shipped to the Wellcome Sanger Institute for library preparation and sequencing. Libraries were constructed using the NEBNext^®^ Ultra II^™^ RNA library prep kit on an Agilent Bravo WS automation system followed by 14 cycles of PCR using KAPA HiFi HotStart DNA polymerase. Libraries were then pooled in equimolar amounts and 75bp paired end (PE) reads were generated on the Illumina HiSeq v4 according to the manufacturer’s standard protocol (∼ 35 million PE reads per sample).

FASTQ files were quality assessed using FASTQC, reads were aligned to the human transcriptome (Ensembl, release 98) using bowtie2 (v2.2.7) (89) and after alignment globin reads were discarded. DESeq2 (39) was used for all differential gene expression analysis; differentially expressed genes were classified as those with an adj p < 0.05 and an absolute fold change > 1.5. To visualise differentially expressed genes in the heatmap in figure 8A all genes with multiple differentially expressed transcripts were filtered to retain the transcript with the lowest adj p value - this was taken forward to estimate fold change.

Gene ontology analysis was performed on lists of differentially expressed genes in Cytoscape (version 3.8.0 using Java 11.0.9.1 in Ubuntu) using the ClueGO plugin (version 2.5.7) (40, 41). The ontologies GO Biological Process-EBI-uniprot-GOA (updated 08.05.2020) and GO Molecular Function-EBI-uniprot-GOA (updated 08.05.2020) were both used. ClueGO networks were constructed using GO term levels 5-11, GO fusion = true and a lower cut-off of 3 genes (or 5% associated genes). The lower bound for connecting GO terms with shared genes was set at a kappa score of 0.4.

### CyTOF sample acquisition

Whole blood samples were taken at baseline, C10, diagnosis and T6 and stabilised in whole blood preservation buffer (Cytodelics AB) within 30 minutes of blood draw. Preserved samples were stored at -80°C. Samples were thawed in a water bath at 37°C and then fixed and red cells lysed using the whole blood preservation kit (Cytodelics AB) according to the manufacturer’s instructions. Fixed samples were washed, permeabilised and barcoded using the Cell-ID 20-Plex Pd Barcoding Kit (Fluidigm). Barcoded samples were then pooled and counted before resuspending in cell staining buffer (Fluidigm) at 40 × 10^6^ cells ml^-1^. An equal volume of freshly prepared surface antibody cocktail (supplementary file 2) was added for 30 minutes and incubated at room temperature under gentle agitation. After washing, samples were resuspended in nuclear antigen staining buffer for 30 minutes at room temperature under gentle agitation. Samples were then washed in nuclear antigen staining perm buffer (Fluidigm) before antibodies for nuclear targets were added and incubated for a further 45 minutes. After another round of washing cells were fixed using 1.6% paraformaldehyde in PBS for 10 minutes and then finally resuspended in fix and perm buffer (Fluidigm) and 72.5 nM Cell-ID™ 191Ir/193Ir intercalator (Fluidigm) at a concentration of 2 x10^6^ cells ml^-1^. Cells were incubated overnight at 4°C.

Sample acquisition was performed on a freshly tuned Helios mass cytometer (Fluidigm) using the WB injector and acquired with 10% normalisation beads (140Ce, 151Eu, 165Ho, and 175Lu, all Fluidigm). Both staining and sample acquisition were carried out in two batches (all time-points for 3 volunteers per batch). On each acquisition day, pooled cells were counted again before removing an aliquot of 2 × 10^6^ cells; aliquots were washed twice in cell staining buffer and resuspended in 1ml cell acquisition solution (Fluidigm). Each aliquot was acquired completely before washing and processing the next aliquot until all pooled samples had been acquired. Cells were acquired at a rate of 300-500 events per second.

### CyTOF data analysis

FCS files were generated using CyTOF software (version 6.7, Fluidigm) followed by normalisation (90) and debarcoding (91) using the *CATALYST* workflow described in (92). Single-stained beads were used for compensation (using non-negative linear least squares regression (93)) and FCS files were gated in the Cytobank web portal (Beckmann Coulter) to exclude normalisation beads and doublets. Singlet T cells (CD45^+^CD3^+^CD19^-^) were taken forward for analysis. Intensity distributions of each channel were inspected to remove channels of low variance (CD14, Tim3, Integrin β7, CD56, CD16, CD49d, CD103, CXCR5). Of note, low variance in these channels does not necessarily reflect uniform or absent expression of these markers, but could also be due to inefficient staining of fixed samples using these antibody clones. The remaining 28 markers were used for both UMAP projections and FlowSOM clustering.

UMAP (46) creates low-dimensional projections of high-dimensional data. Here cells were grouped according to marker expression intensity and embedded in a 2D plane such that phenotypic similarity within and between populations is preserved in the Euclidean distance of the projection. We used its R implementation in the *scater* package (94), which in turn relies on *uwot* (github.com/jlmelville/uwot). Features were scaled to unit variance and the 15 nearest neighbours were considered for embedding. UMAP coordinates were then exported for visualisation using *ggplot2*.

FlowSOM (47) uses self-organising maps (SOM) to efficiently categorise cytometry data into non-overlapping cell populations. Clustering was performed with a target cluster number of 100 and metaclustering with a target cluster number of 45. This approach purposefully overclustered the data to resolve potentially small subsets, a trade-off that can split phenotypically similar cells into more than one population (95). Overclustering was addressed by manual inspection of all clusters and merging of phenotypically similar populations. After manual merging, each T cell was classified into one of 34 unique clusters. Names were assigned manually using activation, lineage and memory markers to broadly categorise each T cell cluster (see supplementary figure 2); when more than one cluster was placed into the same category clusters were given an accessory label to highlight their unique phenotype or property (e.g. skin-homing, indicated by the expression of CLA). The R/Bioconductor package *ComplexHeatmap* was used to visualise T cell cluster phenotypes; the arcsine transformed signal intensity of each marker was independently scaled using a 0- 1 transformation across all 34 clusters.

For differential abundance analysis we followed the workflow laid out in (92). FlowSOM cluster cell counts were modelled linearly with time-point as a dependent categorical variable and volunteer as a fixed effect using the *diffcyt* (49) implementation of edgeR (48). The edgeR functions automatically normalise cluster counts for the total number of cells and improve statistical power by sharing information on cluster count variance between clusters. Pairwise comparisons were performed relative to baseline, and clusters with an FDR < 0.05 and absolute fold change > 2 were deemed to vary significantly through time.

### Pearson correlation analysis

The fold change of each T cell cluster and plasma analyte was calculated using raw cluster percentages or plasma concentrations, respectively. For each feature, these were calculated at diagnosis or T6 (relative to baseline) according to their largest absolute fold change. All data were log2 transformed and Pearson correlation was performed using the cor function from the *stats* R package. Correlation coefficients were then used for hierarchical clustering by Euclidean distance using *ComplexHeatmap*.

### Comparing the immune response between *P. vivax* and *P. falciparum*

For the comparison of *P. vivax* and *P. falciparum* data from a previously conducted clinical trial (VAC063C) were used; the VAC063C trial is reported in full elsewhere (Muñoz Sandoval *et al*., in preparation). In brief, three healthy malaria-naive adult volunteers were inoculated with *P. falciparum* (clone 3D7) by routine blood challenge (approx. 800 parasites per volunteer) (96). Starting one day after infection volunteers attended clinic for assessment and blood sampling every 12-hours, and parasite density was measured in real time by qPCR (target gene = 18S ribosomal RNA). At diagnosis (asymptomatic with parasitaemia above 10,000 parasites ml^-1^ or symptomatic with parasitaemia above 5000 parasites ml^-1^) volunteers were treated with either Riamet or Malarone. VAC063C was sponsored by the University of Oxford, received ethical approval from UK NHS Research Ethics Service (South Central - Oxford A reference 18/SC/0521) and was registered on ClinicalTrials.gov (NCT03906474).

Blood for immunological analyses was collected in EDTA tubes by venepuncture at baseline, during infection, diagnosis, T6 and 45-days post-challenge (memory phase). To undertake whole blood RNA-sequencing samples were processed exactly as described above for *P. vivax* CHMI. Libraries were then prepared using the TruSeq Stranded mRNA library prep kit (Illumina) and sequenced on the NovaSeq 6,000 Illumina platform to yield 50 bp paired end (PE) reads. Data processing and analysis was carried out analogously to VAC069A to identify differentially expressed genes at diagnosis and T6 (versus baseline). ClueGO networks were constructed by combining separate lists of differentially expressed genes from volunteers infected with *P. vivax* and *P. falciparum*. For each GO term information on what fraction of associated genes were derived from the list of DEG in vivax or falciparum malaria was retained. Any GO term containing > 65% associated genes from a single volunteer cohort was considered to be enriched in that infection model, otherwise GO terms were considered to be shared.

For CyTOF analysis whole blood samples were preserved, processed and acquired on a Helios mass cytometer exactly as described for *P. vivax* CHMI. Data processing was also performed analogously to VAC069A and FlowSOM was used to place each T cell into a phenotypically unique cluster. Note that clustering was performed independently on the VAC069A and VAC063C datasets. For each volunteer, the proportion of activated CD4^+^ T cells and regulatory T cells was then calculated by summing the frequency of all clusters with a CD38^hi^Bcl2^lo^ phenotype.

## Data Availability

RNA-sequencing data from VAC069A have been deposited in the European Genome-phenome Archive (EGA) and are accessible through accession number EGAS00001003847. RNA-seq data from VAC063C have been deposited in NCBIs Gene Expression Omnibus and are accessible through accession number GSE172450. CyTOF (mass cytometry) data are available at flowrepository.org and can be accessed through experiment number FR-FCM-Z3HA.

https://flowrepository.org/id/FR-FCM-Z3HA

https://www.ebi.ac.uk/ega/studies/EGAS00001003847

https://www.ncbi.nlm.nih.gov/geo/query/acc.cgi?acc=GSE172450

## Data access

RNA-sequencing data from VAC069A have been deposited in the European Genome-phenome Archive (EGA) and are accessible through accession number EGAS00001003847. RNA-seq data from VAC063C have been deposited in NCBI’s Gene Expression Omnibus and are accessible through accession number GSE172450. CyTOF (mass cytometry) data are available at flowrepository.org and can be accessed through experiment number FR-FCM-Z3HA.

## Acknowledgements

The authors would like to thank Mandy Sanders and the DNA Pipelines team at the Wellcome Sanger Institute for their support with RNA-seq library preparation and data production. We would also like to thank the VAC069A and VAC063C clinical and laboratory study teams for assistance (University of Oxford), and all of the clinical trial volunteers who participated in the studies.

## Declaration of interests

The authors declare no competing interests.

**Supplementary figure 1.**
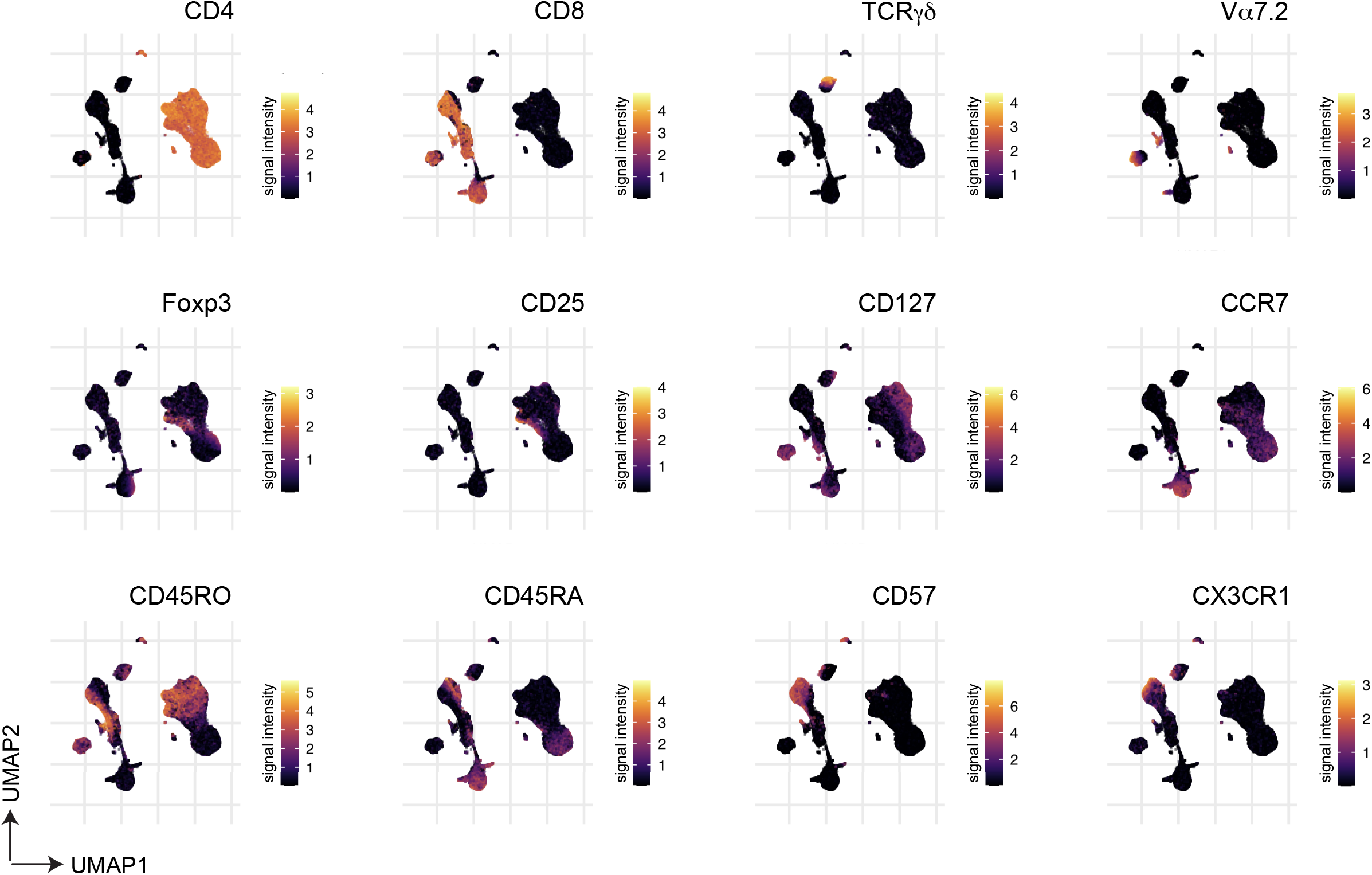
Expression of lineage and memory markers across the UMAP projection. Data shown at T6; used to pinpoint the location of each major T cell subset. The arcsine transformed signal intensity is plotted for each marker.

**Supplementary figure 2.**
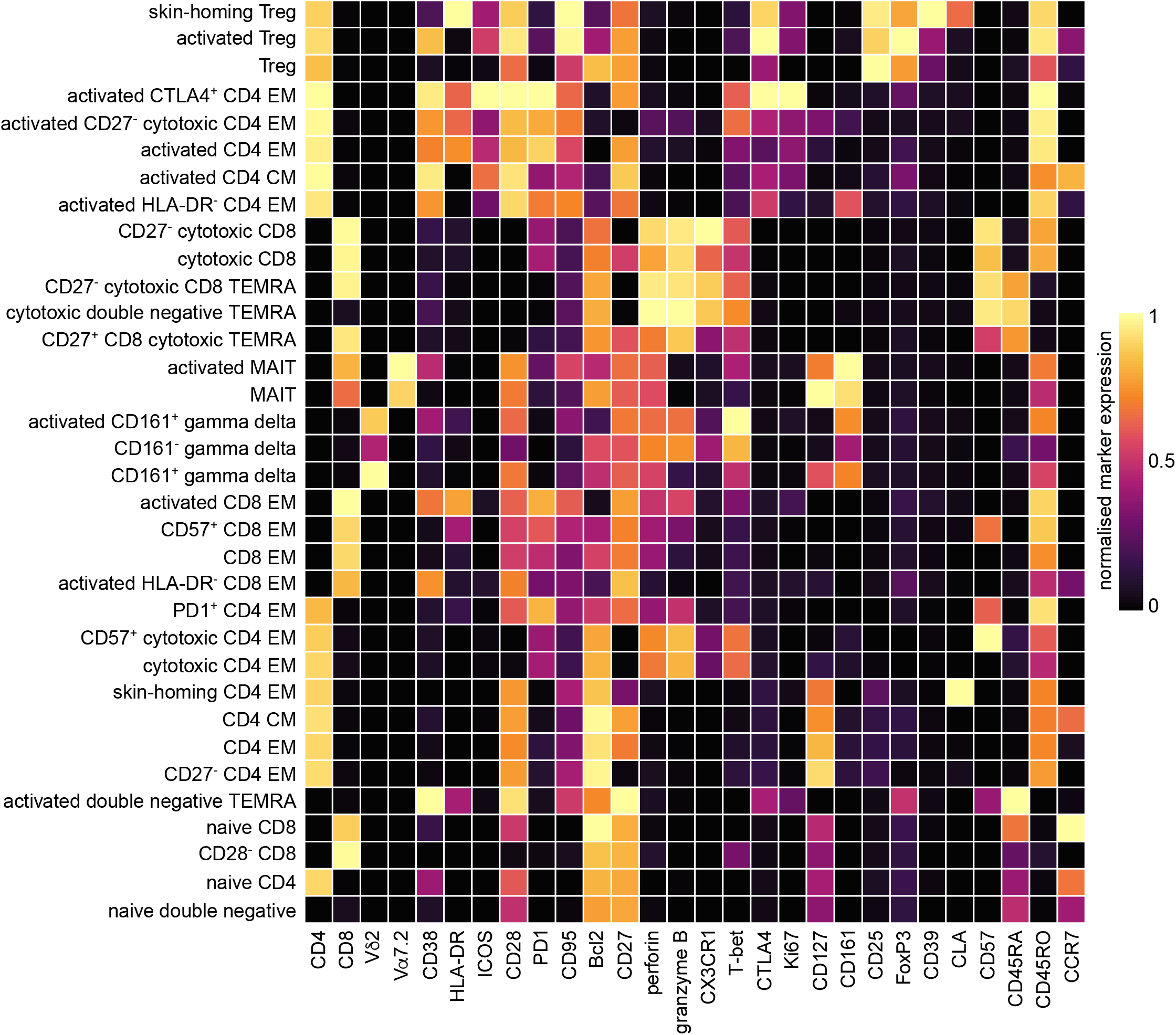
Phenotype of each unique T cell cluster. Heatmap showing normalised median expression values of all markers used for clustering in each of the 34 T cell clusters. Names were assigned manually using activation, lineage and memory markers to broadly categorise each T cell cluster; when more than one cluster was placed into the same category (e.g. activated CD4 EM) clusters were given an accessory label to highlight their unique phenotype or property (e.g. skin-homing, indicated by the expression of CLA).

**Supplementary figure 3.**
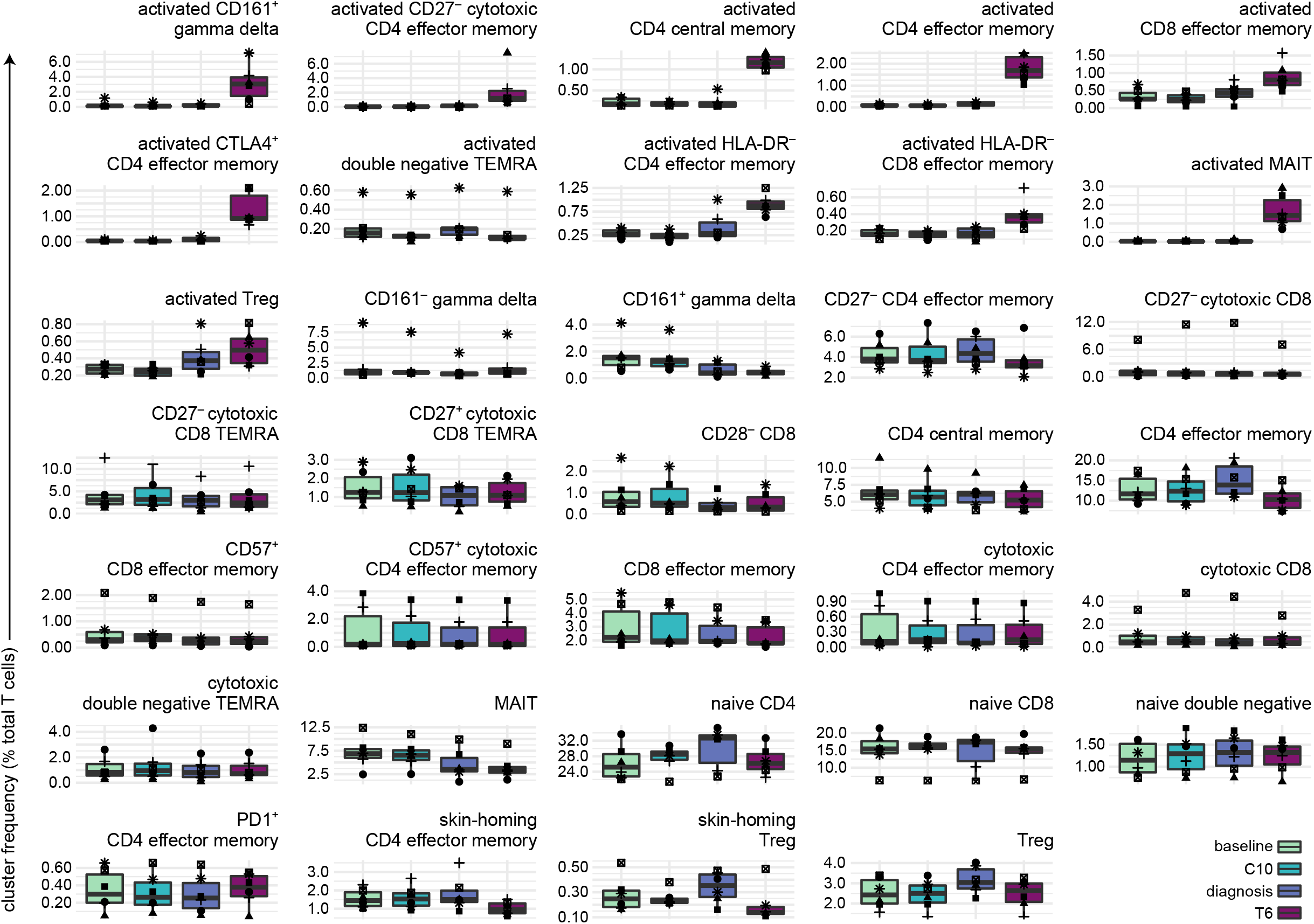
Frequency of each T cell cluster during and after infection. Every unique T cell cluster is shown as the proportion of total T cells at each time-point.

**Supplementary figure 4.**
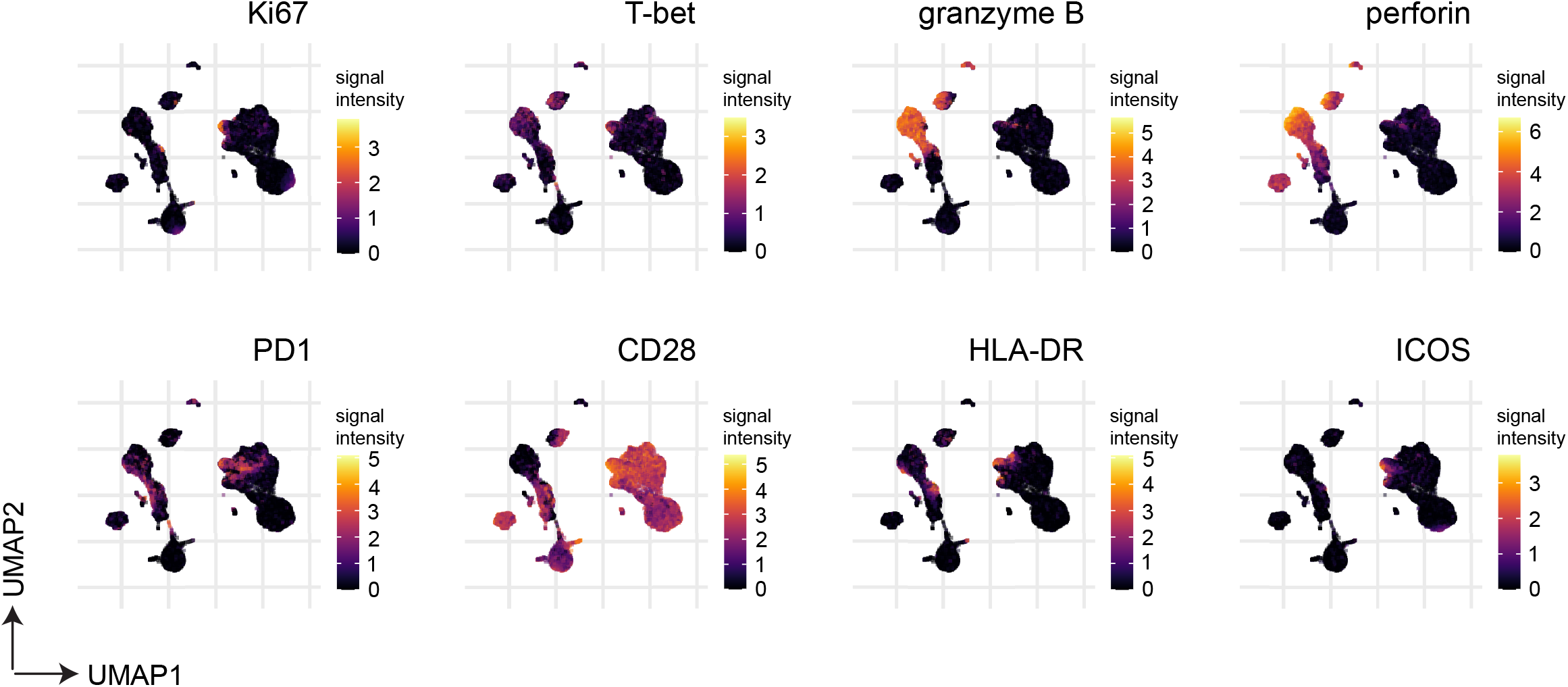
Expression of activation, proliferation and differentiation markers across the UMAP projection. Data shown at T6; allows for a direct comparison of expression levels between activated CD4^+^ T cells and all other T cell subsets. The arcsine transformed signal intensity is plotted for each marker.

## Supplementary Files

**Supplementary file 1**. Demographics of volunteers infected with *Plasmodium vivax* (PvW1) by blood challenge; includes genetic and non-genetic variables known to influence human immune variation *in vitro*.

**Supplementary file 2**. Mass cytometry antibody panel for T cell fate and function in vivax malaria; includes information on the antibody clone and heavy metal conjugate.

